# The associations between Screen Time, Screen Content, and ADHD risk based on the evidence of 41494 children from Longhua district, Shenzhen, China

**DOI:** 10.1101/2024.10.12.24315388

**Authors:** Jian-Bo Wu, Yanni Yang, Qiang Zhou, Jiemin Li, Wei-Kang Yang, Xiaona Yin, Shuang-Yan Qiu, Jingyu Zhang, Minghui Meng, Jian-hui Chen, Zhaodi Chen

**Author notes:** These authors also contributed equally to this work (Jian-hui Chen, Zhaodi Chen).

## Abstract

**Objective:** This study investigates the relationship between screen time, screen content, and the risk of Attention Deficit Hyperactivity Disorder (ADHD) using data from a large sample. Specifically, it examines how different types of screen content (such as educational videos, cartoon videos, and interactive videos) are associated with the risk of ADHD. The aim is to offer a scientific foundation for the rational management of children’s screen time and screen content.

**Methods:** We collected data through a questionnaire survey involving a study population of 41,494 children from Longhua District, Shenzhen City, China. The questionnaire recorded the daily screen time and the type of content viewed by the children at ages 1-3 years and assessed their risk of ADHD using the Strengths and Difficulties Questionnaire (SDQ) at ages 4-6 years. Hierarchical logistic regression analysis, controlling for confounding factors, was employed to explore the associations between screen time, screen content, and ADHD risk.

**Results:** In the total sample, 6.7% of the participants had screen time exceeding 60 minutes per day, with educational videos predominant type (63.4%). 16.5% of the participants were identified as being at risk for ADHD. Statistically significant positive associations with ADHD were observed across all categories of screen time (*P*<0.001). Moreover, as screen time increased, the risk of ADHD also rose (*OR*_1∼60 mins/d_=1.627, *95%CI*=1.460∼1.813; *OR*_61∼120 mins/d_=2.838, *95%CI*=2.469∼3.261; *OR*_>120 mins/d_=3.687, *95%CI*=2.835∼4.796).

Significant positive associations with ADHD were observed across all categories of screen time in the educational videos and cartoon videos. For the educational videos group, the odds ratios were as follows: *OR*_1-60 mins/day_=1.683 (*95% CI*=1.481-1.913), *OR*_61-120 mins/day_=3.193 (*95% CI*=2.658-3.835), and *OR*_>120 mins/day_=3.070 (*95% CI*=2.017-4.673). For the cartoon videos group, the odds ratios were: *OR*_1-60 mins/day_=1.603 (*95% CI*=1.290-1.991), *OR*_61-120 mins/day_=2.758 (*95% CI*=2.156-3.529), and *OR*_>120 mins/day_=4.097 (*95% CI*=2.760-6.081).

However, no significant associations with ADHD risk were found for any category of screen time in the interactive videos group (*OR*_1∼60 mins/d_=0.744, *95%CI*=0.361∼1.534; *OR*_61∼120 mins/d_=0.680, *95%CI*=0.296∼1.560; *OR*_>120 mins/d_=1.678, *95%CI*=0.593∼4.748).

**Conclusion:** As screen time increases, the risk of ADHD also rises. Both educational videos and cartoon videos show a positive correlation between screen time and ADHD risk. However, no significant association was found between screen time and ADHD risk when it came to interactive videos. This study underscores the importance of reasonably managing children’s screen time, particularly the time spent watching educational and cartoon videos.

## Introduction

In recent years, the widespread adoption of electronic devices has significantly increased children’s screen time, raising widespread societal concern about the impact of electronic screens on children’s mental health[1, 2]. Most existing research focuses on the association between screen time and the risk of ADHD, highlighting its potential impact on children’s mental health development[3, 4]. However, these studies often overlook the fact that different types of screen content may have distinct effects on children’s mental health[5].

ADHD, a common neurodevelopmental disorder, is characterized by core symptoms such as inattention, hyperactivity, and impulsivity, which pose challenges to children’s learning, social interaction, and emotional regulation abilities[6, 7]. Although the exact pathophysiology of ADHD is not fully understood, environmental factors-including lifestyle factors-are recognized as playing a significant role in its development[8]. Among these factors, the relationship between screen time and ADHD risk has been a hot topic of research in recent years. Howere, studies specifically investigating the association between different type of screen content and ADHD risk remain scarce.

Existing studies on the link between screen time and hyperactive behavior generally indicate that excessive screen time may indirectly increase the risk of ADHD by affecting children’s sleep, physical activity, and social interaction[9, 10]. However, these studies often fail to explore the differential impacts of various types of screen contents on children’s mental health in depth[11, 12]. Educational and cartoon videos, despite their perceived educational value, are primarily one-way information inputs that lack interaction. Their rapidly changing visuals and intense sensory stimulation may adversely affect children’s ability to concentrate and self-regulate[13]. In contrast, interactive videos may more effectively promote children’s social interaction and cognitive development, though their specific impact on ADHD risk requires further investigation.

This study leverages the extensive data resources from the Longhua Child Cohort Study (LCCS) in Shenzhen to comprehensively assess the relationship between children’s screen time, different types of screen content, and the risk of ADHD. The aim is to provide scientific evidence to guide parents and educators in reasonably managing children’s screen time and optimizing screen content selection, thereby effectively reducing the risk of ADHD in children.

## Methods

### Study design and participants

This study was conducted in accordance with the Declaration of Helsinki. The data for this project were sourced from the 2021 survey of the LCCS. The LCCS was a large-scale epidemiological survey conducted in Longhua District, Shenzhen, China, aiming to assess the impact of children’s lifestyle habits on early psychological and behavioral development of preschoolers.

The research project was conducted across 250 kindergartens in the Longhua District of Shenzhen in 2021. From 8th October to 23th November 2021, the project was publicized to the parents of kindergarten children, encouraging their participation. After obtaining parental consent, informed consent forms were signed by the parents, and a questionnaire survey was conducted. A total of 59,600 questionnaires were distributed, and 56,740 were returned, yielding a response rate of 95.2%. After excluding 15,246 questionnaires with incomplete information, the final sample size was 41,494. This study was approved by the Ethics Committee of Longhua Maternal and Child Health Hospital (Ethics approval No. 2016102501).

### Data collection

The questionnaire collected information on family demographic characteristics, daily screen time, and the types of programs viewed during screen time by children when they were at the age of 1-3 years old. Additionally, it assessed the risk of ADHD using the Strengths and Difficulties Questionnaire (SDQ) for them at the age of 4-7 years old. All participants had signed the Human Ethics and Consent to Participate forms and agreed to be involved in this study.

### Measurement of screen time (major exposure variables) and category of

“Screen time” was defined as time spent looking at screens such as phones, TVS, tablets or desktop computers, game consoles, as reported by the children’s parents. We chose to collect information on screen time for children aged 4 to 7. An ordinal categorical survey was conducted to assess screen time, and a nominal categorical survey was conducted to evaluate the types of programs viewed during screen time (Table 1).

**Table 1.** Questions and options regarding the screen time and program of the screen time.

| No. | Questions | Options |
| --- | --- | --- |
| Q1 | How long was the screen time of your child/children at his/her/their age of 1-3 years old per day? | 0 mins/d<br>1~60 mins/d<br>61~120 mins/d<br>>120 mins/d |
| Q2 | What types of programs did your child/children primarily view during screen time at his/her/their age of 1-3 years old? | Educational videos<br>Cartoon videos<br>Interactive videos |

\*Educational videos : (with explicit prosocial or cognitive components [eg, Sesame Street and QiaoHu, a similar program in China])
\*Cartoon videos : (eg, Thomas & Friends and Super Flying Man)
\*Interactive videos (social media or interactive game)

**Table 2.** ADHD scores and risk of the Strengths and Difficulties Questionnaire (SDQ)

| Category | ADHD scores (Mean±SD) | ADHD risk (%) |
| --- | --- | --- |
| Total | 3.69±1.93 | 41494 (100.0) |
| Normal | 3.09±1.45 | 34645 (83.5) |
| Edge value | 6.00±0.00 | 3737 (9.0) |
| Abnormal | 7.60±0.84 | 3112 (7.5) |

### Measurement of ADHD risk

In this study, the Strengths and Difficulties Questionnaire (SDQ) was used to assess the risk of ADHD in children. The SDQ, developed by American psychologist Goodman in 1997, is a concise behavioral screening scale[14]. In 2005, China established norms for the Chinese population[15]. The scale consists of 25 items, covering 5 dimensions: emotional symptoms, conduct problems, ADHD symptoms, peer problems, and prosocial behavior. Items on the SDQ are rated on a scale from 0 to 2, with 0 indicating no agreement, 1 indicating partial agreement, and 2 indicating perfect agreement. The total score for ADHD symptoms ranges from 0 to 5 for normal, 6 for borderline, and 7 to 10 for abnormal. Based on these scores, participants can be categorized into a normal group (≤5) and an ADHD risk group (≥6). The scale demonstrated good reliability, with a Cronbach’s α coefficient of 0.749[16, 17].

### Covariates

The following confounding covariates were included in the analysis: child’s gender, age, parental marital status, parents’ educational attainment, household monthly income, single-child status, and whether the content of screen time programs was discussed with the child.

### Statistical analysis

Descriptive statistics were used to characterize the study population. Mean ± standard deviation (SD) and sample number (percentage) were presented for continuous and categorical variables, respectively. A chi-square test was used to compare differences in screen time, types of programs viewed during screen time, and covariate variables among ADHD risk groups. Logistic regression analysis was employed to explore the association between screen time and ADHD risk.

## Results

### Participants sociodemographic characteristics and differences in screen time, types of programs viewed during screen time, and covariate variables among ADHD risk groups

Participants’ SDQ scores and associated ADHD risk levels are presented in Table 1. In the total sample, we found that 7.5% of the participants exhibited abnormal levels of ADHD symptoms (defined as a score of 7-10). Additionally, 9.0% of the participants were on the borderline for ADHD symptoms (defined as a score of 6).

Overall, 16.5% of the participants were at risk for ADHD.

Participants ‘ demographics and characteristics are summarized in Table 3. A total of 41,494 children (22,113 boys [53.3%] and 19,381 girls [46.7%]; mean [SD] age, 5.13 ±0.67 years old) completed the questionnaire. The risk of ADHD was higher in boys compared to girls(18.9% vs. 13.7%, *χ*^*2*^=201.855, *P*<0.001).

**Table 3.**
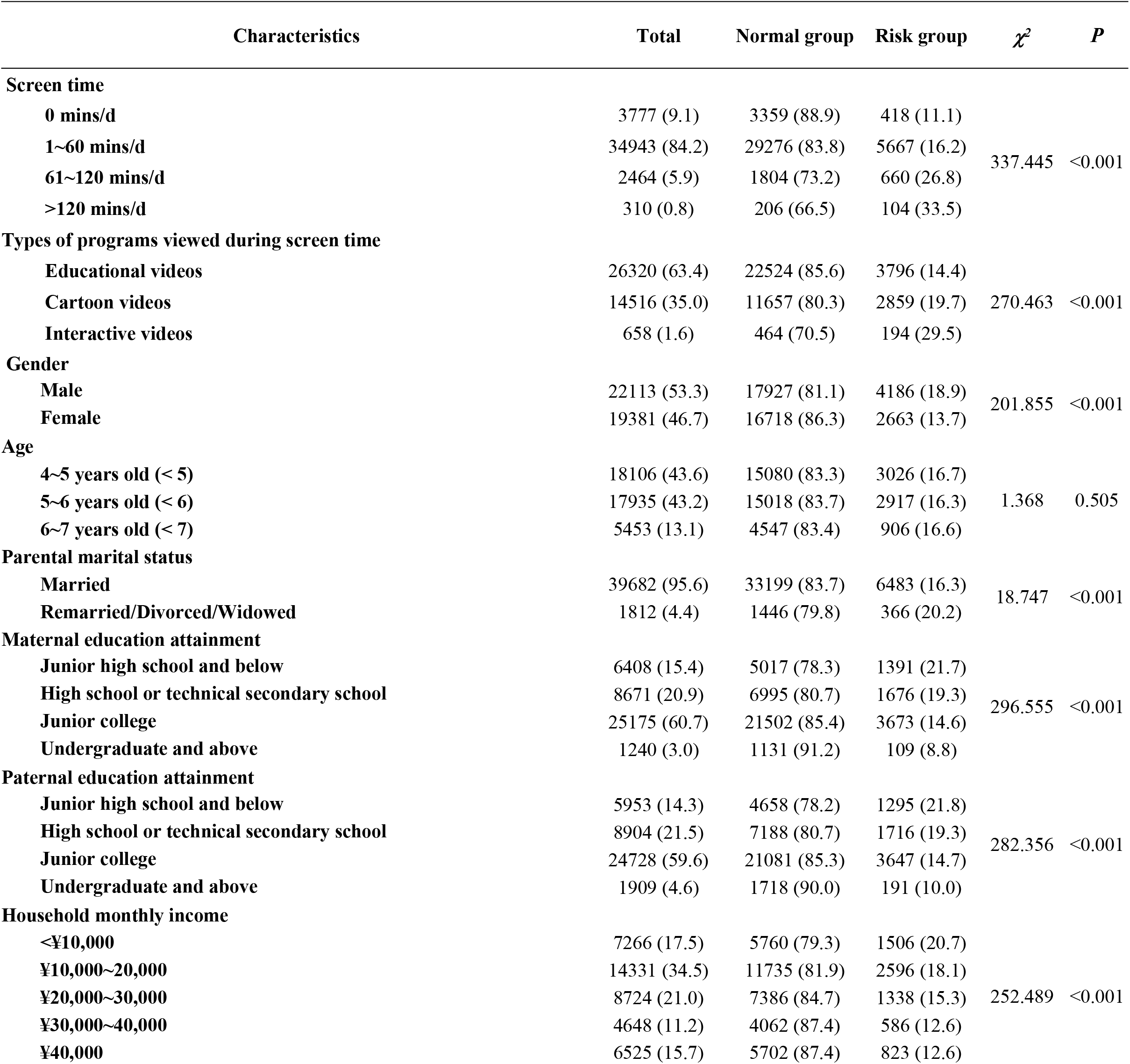
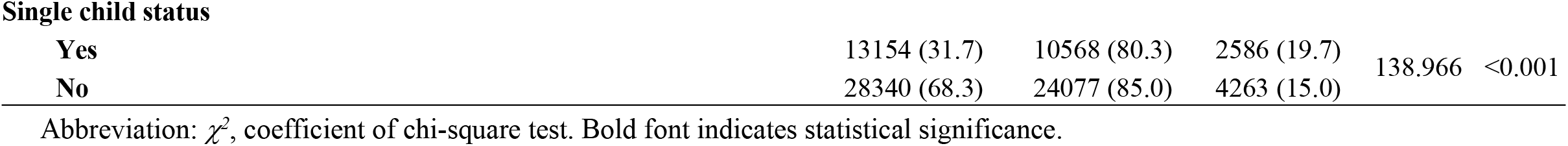
Participants sociodemographic characteristics and differences in screen time, types of programs viewed during screen time, and covariate variables among ADHD risk groups(N=41,494)

### Relationship between screen time and ADHD risk

We performed a logistic regression analysis to investigate the associations between screen time and ADHD risk (Table 4). Statistically significant positive associations with ADHD were observed across all categories of screen time (*β*_1∼60 mins/d_ = 0.487, *β*_61∼120 mins/d_ = 1.043, *β*_>120 mins/d_ = 1.305, *P*<0.001). Additionally, as screen time increased, the risk of ADHD also rose (*OR*_1∼60 mins/d_=1.627, *95%CI*=1.460∼1.813; *OR*_61∼120 mins/d_=2.838, *95%CI*=2.469∼3.261; *OR*_>120 mins/d_=3.687, *95%CI*=2.835∼4.796).

**Table 4.** Relationship between screen time and ADHD risk (N=41,494)

| Screen time | $\beta$ | OR (95%CI) | P |
| --- | --- | --- | --- |
| 0 mins/d |  | Ref. |  |
| 1~60 mins/d | 0.487 | 1.627 (1.460~1.813) | <0.001 |
| 61~120 mins/d | 1.043 | 2.838 (2.469~3.261) | <0.001 |
| >120 mins/d | 1.305 | 3.687 (2.835~4.796) | <0.001 |

Screen time was transferred into dummy variables. Abbreviation: $\beta$ , coefficient of Logistic Regression Analysis with adjustment for age, gender, parental marital status, maternal and paternal education attainment, household monthly income, single child status, types of programs viewed during screen time and discuss the content of screen time program with the child. OR, odds ratio of Logistic Regression Analysis. 95%CI, 95% confidence interval of OR of Logistic Regression Analysis. Bold font indicates statistical significance.

### Relationship between screen time and ADHD risk in different types of programs viewed during screen time

We conducted a stratified logistic regression analysis to further investigate the associations between screen time and ADHD risk across different types of programs viewed during screen time (Table 5). Statistically significant positive associations with ADHD were observed across all categories of screen time in the educational videos and cartoon videos groups. (education videos group [*OR*_1∼60 mins/d_=1.683, *95%CI*=1.481∼1.913; *OR*_61∼120 mins/d_=3.193, *95%CI*=2.658∼3.835; *OR*_>120 mins/d_=3.070, *95%CI*=2.017∼4.673]; Cartoon group [*OR*_1∼60 mins/d_=1.603, *95%CI*=1.290∼1.991; *OR*_61∼120 mins/d_=2.758, *95%CI*=2.156∼3.529; *OR*_>120 mins/d_=4.097, *95%CI*=2.760∼6.081]) However, no category of screen time was significantly associated with ADHD risk in the social software group (*OR*_1∼60 mins/d_=0.744, *95%CI*=0.361∼1.534; *OR*_61∼120 mins/d_=0.680, *95%CI*=0.296∼1.560; *OR*_>120 mins/d_=1.678, *95%CI*=0.593∼4.748).

**Table 5.** Relationship between screen time and ADHD risk in different types of programs viewed during screen time (N=41,494) Screen time was transferred into dummy variables. Abbreviation: *OR*(*95%CI*), odds ratio(95% confidence interval of *OR*)of Logistic Regression Analysis with adjustment for age, gender, parental marital status, maternal and paternal education attainment, household monthly income, single child status and discuss the content of screen time program with the child. Bold font indicates statistical significance.

| Type of content viewed during screen time | Educational videos (n=26320) | Cartoon videos (n=14516) | Interactive videos (n=658) |
| --- | --- | --- | --- |
| 0 mins/d |  | Ref. |  |
| 1~60 mins/d | 1.683 (1.481~1.913) | 1.603 (1.290~1.991) | 0.744 (0.361~1.534) |
| 61~120 mins/d | 3.193 (2.658~3.835) | 2.758 (2.156~3.529) | 0.680 (0.296~1.560) |
| >120 mins/d | 3.070 (2.017~4.673) | 4.097 (2.760~6.081) | 1.678 (0.593~4.748) |

## Discussion

### The Relationship between Screen Time and ADHD Risk

In this study, we observed a significant positive correlation between screen time and the risk of ADHD among children. This association may be attributed to the combined effects of multiple mechanisms. Firstly, the use of screen devices, especially before bedtime, may disrupt children’s sleep patterns by inhibiting the secretion of melatonin, thereby affecting their attention and emotional regulation abilities[18-20]. Secondly, the fast-paced and highly stimulating content on screens may overstimulate children’s attention systems, impacting their cognitive and attentional development[21, 22]. Additionally, increased screen time often comes at the expense of physical activity, which has a benificial effect on improving children’s attention and reducing hyperactive behaviors[23, 24]. Furthermore, excessive screen time may reduce children’s social interactions with peers, and a lack of social skills is associated with the development of ADHD, leading to peer relationship problems and difficulties in school adjustment[25-27]. These findings underscore the importance of reducing children’s screen time to preventing ADHD.

### The Relationship between Screen content and ADHD Risk

Educational videos, as a medium for children to acquire knowledge, do not universally exert positive influences[28-30]. Previous research findings on educational videos have been inconsistent. Some studies suggest that educational videos do not significantly increase the risk of attention deficit hyperactivity disorder (ADHD)[31], while others have found that prolonged exposure to educational videos may adversely affect children’s attentional systems [32]. This study reveals that as the time spent watching educational videos increases, the risk of ADHD among children rises significantly. This may be attributed to the fact that educational videos often contain a wealth of information with rapid scene changes, which can easily overstimulate children’s attentional systems, thereby impairing their self-regulation abilities[33, 34]. Additionally, the lack of interactivity in educational videos may deprive children of opportunities to practice and apply knowledge in real-life situations, impacting their social skills and problem-solving abilities[35, 36].

Cartoon videos, with their vibrant colors and exaggerated movements, are popular among children, but prolonged viewing may also elevate the risk of ADHD. The fast-paced and highly stimulating content in cartoon videos may excessively activate children’s attentional systems[37], leading to decreased attention to the real world and affecting cognitive and emotional development [38]. Moreover, violent or stimulating content in cartoon videos may adversely affect children’s mental health[39, 40], further increasing the risk of ADHD.

This study finds that, unlike educational and cartoon videos, interactive videos do not show a significant association with the risk of ADHD. Biofeedback therapy, used in attention training for children with ADHD[41], leverages interactive videos to train children’s attention. During these sessions, children must adjust their brain electrical activity in real-time while watching interactive videos, thereby enhancing their attention [42, 43]. Interactive videos provide children with a greater sense of participation and feedback opportunities[44, 45], which may contribute to the lack of significant association between interactive videos and ADHD risk. However, this does not imply that children should increase their use of interactive videos indiscriminately.

This study possesses notable strengths, primarily reflected in its large sample size (41,494 children) and meticulous data analysis, allowing for an in-depth investigation into the relationship between screen time and ADHD risk, as well as the specific impacts of various screen contents (educational videos, cartoon videos, and interactive videos). However, this study also has several limitations. Firstly, the cross-sectional design precludes direct inference of causality, necessitating future research to adopt a longitudinal design for further validation of the findings. Secondly, the data relies on parental recall, which may be subject to recall bias. Lastly, despite controlling for multiple confounding factors, there may still be other unrecognized factors influencing the results.

## Conclusion

As screen time increases, so does the risk of ADHD. For both educational and cartoon videos, screen time is positively correlated with ADHD risk. However, no significant association between screen time and ADHD risk was found when using interactive videos. This study underscores the importance of reasonably controlling children’s screen time, particularly the time spent watching educational videos and cartoon videos.

## Data Availability

All relevant data are within the manuscript and its Supporting Information files.

## Acknowledgments

The author would like to express his gratitude to the participants of the study, the investigators of Shenzhen Longhua District Maternal and Child Health Hospital and the kindergarten teachers who participated in the survey.

## Author Contributions

**Conceptualization:** Jian-Bo Wu and Jian-hui Chenki.

**Data curation:** Shuang-Yan Qiu, Jian-Bo Wu and Zhaodi Chen.

**Formal analysis:** Jian-Bo Wu and Jian-hui Chen.

**Funding acquisition:** Zhaodi Chen, Jian-Bo Wu, Jie-Min Li, Qiang Zhou.

**Investigation:** Jingyu Zhang, Jian-Bo Wu and Shuang-Yan Qiu.

**Methodology:** Zhaodi Chen and Jian-hui Chen.

**Project administration:** Jian-Bo Wu, Xiaona Yin and Yanni Yang.

**Resources:** Jian-Bo Wu and Zhaodi Chen.

**Software:** Yanni Yang and Jian-hui Chen.

**Supervision:** Wei-Kang Yang and Minghui Meng.

**Validation:** Jian-Bo Wu and Zhaodi Chen.

**Visualization:** Jian-Bo Wu and Jian-hui Chen.

**Writing–original draft:** Jian-Bo Wu, Jian-hui Chen and Zhaodi Chen.

**Writing–review and editing:** Jian-Bo Wu, Jian-hui Chen and Zhaodi Chen.

## References

[1] Radesky, J.S. and D.A. Christakis, Increased Screen Time: Implications for Early Childhood Development and Behavior. Pediatr Clin North Am, 2016. 63(5): p. 827–39.

[2] VAN Rooij, A.J., et al., The (co-)occurrence of problematic video gaming, substance use, and psychosocial problems in adolescents. J Behav Addict, 2014. 3(3): p. 157–65.

[3] Scholtens, S., A.M. Rydell and F. Yang-Wallentin, ADHD symptoms, academic achievement, self-perception of academic competence and future orientation: a longitudinal study. Scand J Psychol, 2013. 54(3): p. 205–12.

[4] Page, A.S., et al., Children’s screen viewing is related to psychological difficulties irrespective of physical activity. Pediatrics, 2010. 126(5): p. e1011–7.

[5] Beyens, I., P.M. Valkenburg and J.T. Piotrowski, Screen media use and ADHD-related behaviors: Four decades of research. Proc Natl Acad Sci U S A, 2018. 115(40): p. 9875–9881.

[6] Chorozoglou, M., et al., Preschool hyperactivity is associated with long-term economic burden: evidence from a longitudinal health economic analysis of costs incurred across childhood, adolescence and young adulthood. J Child Psychol Psychiatry, 2015. 56(9): p. 966–75.

[7] Tamayo, M.N., et al., Aggressive behavior, emotional, and attention problems across childhood and academic attainment at the end of primary school. Soc Psychiatry Psychiatr Epidemiol, 2021. 56(5): p. 837–846.

[8] Thapar, A. and M. Cooper, Attention deficit hyperactivity disorder. Lancet, 2016. 387(10024): p. 1240–50.

[9] Li, S., et al., The impact of media use on sleep patterns and sleep disorders among school-aged children in China. Sleep, 2007. 30(3): p. 361–7.

[10] Eirich, R., et al., Association of Screen Time With Internalizing and Externalizing Behavior Problems in Children 12 Years or Younger: A Systematic Review and Meta-analysis. JAMA Psychiatry, 2022. 79(5): p. 393–405.

[11] Wu, J.B., et al., Association between screen time and hyperactive behaviors in children under 3 years in China. Front Psychiatry, 2022. 13: p. 977879.

[12] Radesky, J.S., J. Schumacher and B. Zuckerman, Mobile and interactive media use by young children: the good, the bad, and the unknown. Pediatrics, 2015. 135(1): p. 1–3.

[13] Zimmerman, F.J. and D.A. Christakis, Associations between content types of early media exposure and subsequent attentional problems. Pediatrics, 2007. 120(5): p. 986–92.

[14] Dickey, W.C. and S.J. Blumberg, Revisiting the factor structure of the strengths and difficulties questionnaire: United States, 2001. J Am Acad Child Adolesc Psychiatry, 2004. 43(9): p. 1159-67. [15].

[15] Jianhua, K. and D. Yasong, Reliability and validity of “children strengths and difficulties questionnaire” in Shanghai norm. Shanghai Archives of Psychiatry, 2005.

[16] Wu, X., et al., The relationship between screen time, nighttime sleep duration, and behavioural problems in preschool children in China. European Child & Adolescent Psychiatry, 2017. 26(5): p. 541–548.

[17] Du Y, J. Kou and D. Coghill, The validity, reliability and normative scores of the parent, teacher and self report versions of the Strengths and Difficulties Questionnaire in China. Child Adolesc Psychiatry Ment Health, 2008. 2(1): p. 8.

[18] Qiu, S.Y., et al., Relationship between bedtime, nighttime sleep duration, and anxiety symptoms in preschoolers in China. Front Psychol, 2024. 15: p. 1290310.

[19] Leblanc, A.G., et al., Correlates of Total Sedentary Time and Screen Time in 9–11 Year-Old Children around the World: The International Study of Childhood Obesity, Lifestyle and the Environment. PLoS ONE, 2015. 10(6): p. e0129622..

[20] Ke, Y., S. Chen and Y.L.Y. Hong, Associations between socioeconomic status and screen time among children and adolescents in China: A cross-sectional study. plos one, 2023. 18(3).

[21] Zhao, J., et al., Excessive Screen Time and Psychosocial Well-Being: The Mediating Role of Body Mass Index, Sleep Duration, and Parent-Child Interaction. J Pediatr, 2018. 202: p. 157-162.e1.

[22] Nikkelen, S.W., et al., Media use and ADHD-related behaviors in children and adolescents: A meta-analysis. Dev Psychol, 2014. 50(9): p. 2228–41.

[23] Papageorgiou, M., et al., Reduced energy availability: implications for bone health in physically active populations. Eur J Nutr, 2018. 57(3): p. 847–859.

[24] Suchert, V., et al., Relationship between attention-deficit/hyperactivity disorder and sedentary behavior in adolescence: a cross-sectional study. Atten Defic Hyperact Disord, 2017. 9(4): p. 213–218.

[25] Domingues-Montanari, S., Clinical and psychological effects of excessive screen time on children. J Paediatr Child Health, 2017. 53(4): p. 333–338.

[26] Alanko, D., The Health Effects of Video Games in Children and Adolescents. Pediatr Rev, 2023. 44(1): p. 23–32.

[27] Twenge, J.M. and W.K. Campbell, Associations between screen time and lower psychological well-being among children and adolescents: Evidence from a population-based study. Prev Med Rep, 2018. 12: p. 271–283.

[28] Page, A.S., et al., Children’s screen viewing is related to psychological difficulties irrespective of physical activity. Pediatrics, 2010. 126(5): p. e1011–7.

[29] Nereim, C., D. Bickham and M. Rich, A primary care pediatrician’s guide to assessing problematic interactive media use. Curr Opin Pediatr, 2019. 31(4): p. 435–441.

[30] Baydar, N., et al., Effects of an educational television program on preschoolers: Variability in benefits. Journal of Applied Developmental Psychology, 2008. 29(5): p. 349–360.

[31] Wang, H., et al., Types of On-Screen Content and Mental Health in Kindergarten Children. JAMA Pediatr, 2024. 178(2): p. 125–132.

[32] Linebarger and L. D., Infants’ and Toddlers’ Television Viewing and Language Outcomes. American Behavioral Scientist, 2005. 48(5): p. 624–645.

[33] Paulus, F.W., et al., Electronic Media and Early Childhood: A Review. Klin Padiatr, 2021. 233(4): p. 157–172.

[34] Thakkar, R.R., M.M. Garrison and D.A. Christakis, A systematic review for the effects of television viewing by infants and preschoolers. Pediatrics, 2006. 118(5): p. 2025–31.

[35] Linebarger, D.L., et al., Associations between parenting, media use, cumulative risk, and children’s executive functioning. J Dev Behav Pediatr, 2014. 35(6): p. 367–77.

[36] Fitzpatrick, C., T. Barnett and L.S. Pagani, Early exposure to media violence and later child adjustment. J Dev Behav Pediatr, 2012. 33(4): p. 291–7.

[37] Kostyrka-Allchorne, K., et al., Differential effects of film on preschool children’s behaviour dependent on editing pace. Acta Paediatrica, 2017. 106.

[38] Verlinden, M., et al., Television viewing through ages 2-5 years and bullying involvement in early elementary school. BMC Public Health, 2014. 14: p. 157.

[39] Christakis, D.A. and F.J. Zimmerman, Violent television viewing during preschool is associated with antisocial behavior during school age. Pediatrics, 2007. 120(5): p. 993–9.

[40] Gentile, D.A. and W. Stone, Violent video game effects on children and adolescents. A review of the literature. Minerva Pediatr, 2005. 57(6): p. 337–58.

[41] Sitaram, R., et al., Closed-loop brain training: the science of neurofeedback. Nature Reviews Neuroscience, 2016. 18(2): p. 86.

[42] Minder, F., et al., Informant-related effects of neurofeedback and cognitive training in children with ADHD including a waiting control phase: a randomized-controlled trial. European Child & Adolescent Psychiatry, 2018.

[43] Van Doren, J., et al., Theta/beta neurofeedback in children with ADHD: Feasibility of a short-term setting and plasticity effects. International Journal of Psychophysiology, 2016: p. 80.

[44] Guo Bingxin et al., Comparison of the effects of EEG biofeedback in the treatment of different subtypes of attention deficit hyperactivity disorder. Chinese Journal of Behavioral Medicine and Brain Sciences, 2021. 30(7): p. 7.

[45] Angelidis, A., et al., Frontal EEG theta/beta ratio as an electrophysiological marker for attentional control and its test-retest reliability. Biological Psychology, 2016. 121(Pt A): p. 49–52.

